# EARLY DETECTION OF COVID-19

**DOI:** 10.1101/2020.04.04.20053363

**Authors:** Hafiz Abdul Sattar Hashmi, Hafiz Muhammad Asif

## Abstract

Since SARS-Cov-2 epidemic appeared in Wuhan China, it became challenge for health authorities to counter Covid-19 epidemic. Early evaluation of suspects, screening for Covid-19 and management posed challenge to health authorities especially in developing countries which were not ready to cope with it. Early mild symptoms appeared during course of disease provide a chance to early detect Covid-19. We use retrospective methodology to collect available data on early sign and symptoms of Covid-19 through accessing World Health Organization (WHO) webpages, New England Journal of Medicine (NEJM), Nature Journal, Journal of American Medical Association (JAMA) network, British Medical Journal (BJM), Lancet and other world renowned journal publications to establish a relationship of early symptoms for detection of Covid-19. Data of 2707 Covid-19 laboratory confirmed cases was collected and analyzed for early signs. Available data was categorized into physical and blood biomarkers. This categorized data was assessed for scoring early detection of Covid-19 by scoring Hashmi-Asif Covid-19 formula. Each characteristic was given a score in Hashmi-Asif Covid-19 chart with maximum score of 28. Scoring 16 on chart means Covid-19 will fully develop in near future. Correlation of each sign and symptoms with development of Covid-19 in formula showed significant correlation assessed by Pearson correlation and Spearman Correlation coefficient (rho) showed significant correlation of development of Covid-19 with fever 64.11% (P=0.001), cough 65% and dry mucus 19.67% equally sensitive (P=0.000), leukopenia 19.06% (P=0.006), lymphopenia 52.93% (P=0.005), thrombopenia 19.1% (P=0.013), elevated Aspartate aminotransferase 12.79% (P=0.007) and elevated Alanine aminotransferase 11.34% (P=0.006). Chart can sense Covid-19 progression 72-96 hours earlier compared to usual course of disease and detection by standard method. Chart for early detection provides early quarantine decision to reduce disease spread and give ample time for intervening disease progression to reduce morbidity time due to Covid-19.

## Background

Since SARS-Cov-2 epidemic appeared in Wuhan China, it became challenge for health authorities to counter Covid-19 epidemic. Early evaluation of suspects, screening for Covid-19 and management posed challenge to health authorities especially in developing countries which were not ready to cope with it. Health care providers faced challenge to initially interpret Covid-19 profiles by early symptoms because course of disease for carriers, infected and complicated profiles were not known previously and required necessarily evaluation of severity of cases presenting during epidemic.^1^ Developing world faced several glitches due to scarcity of diagnostic materials, expensive diagnostic procedures, pre-requisite of highly skilled and trained health staff. Since, developing countries were not ready to cope with Covid-19 consequence to endemic spread. Na Zhu et al, firstly cultured SARS-Cov-2 on cell line and done genetic sequencing and SARS-Cov-2 was numbered seven in family of corona viruses.^2^ Novel Coronavirus infection started with zoonotic contact to humans and cause respiratory symptoms like of pneumonia. On detailed study of characteristics of novel coronavirus given name SARS-Cov-2 because of appearance of similar symptoms of Acute Respiratory Syndrome caused by previously known coronaviruses.^2^ Morens et al, described SARS-Cov-2 has longer incubation period as compared to influenza viruses. Delayed onset of symptoms may provide a chance for early detection and intervention during pathogenesis of Covid-19.^3^ Before severe attack of SARS-Cov-2 to cause Covid-19 mild symptoms appearance holds very important information about the pathogenesis of Covid-19. Li Q et al, provides information on mean incubation period of 5.2 days for Covid-19 symptoms to appear and mean period of 12.5 days for hospitalization from onset of symptoms.^4^ Time period between mild symptoms appearance to critical attack of Covid-19 provides a chance for early detection by tracing common signs and symptoms appeared before severe attack of Covid-19. Early time period of Covid-19 infection extends chance of interjectory interventions. Anthony S. Fauci et al, explained the importance of such delay onset of symptoms and such delay may provide crucial information for intervention to halt Covid-19 which could be possible with better understanding of pathogenesis of covid-19.^5^

## Methods

We used retrospective approach to collect observational data about most common presenting sign and symptoms in reported Covid-19 cases. Renowned Journal websites were assessed with available publication holding important information about the course of disease Covid-19 and containing clinical features and laboratory investigations. Data was also assessed by searching digital tools like Google scholar, and Alta Vista. World Health Organization (WHO) webpages, New England Journal of Medicine (NEJM), Nature Journal, Journal of American Medical Association (JAMA) network, British Medical Journal (BJM) and the Lancet etc. were considered for getting available updated information about the clinical aspects and clinical presentation of Covid-19. Various National Health Departments for Infectious Disease Control center of various developed countries websites and pages offering Covid-19 updates, data were assessed for common presentations made by collected publications for sensing essential common symptoms. Hashmi-Asif Covid-19 Formula was designed based on collected data were scrutinized to most common and easily accessible symptoms which can alter diagnosis of Covid-19 or due to their absence diagnosis could be delayed. Keeping importance of differentiating clinical features were aligned in table form providing a sketch of most common essential symptoms. Keeping frequency of symptoms in relation with Covid-19 diagnosis were categorized into physical and blood biomarkers. Booth were categorized separately and were given separate score according to the severity of symptoms relevant with frequency of the specific symptoms or signs appeared in Covid-19. Minimum and maximum scores were calculated and evaluated for available data collected for provided in Table.1. All data were calculated on score chart to evaluate its efficacy for detecting early signs and symptoms to make an easy decision to hold isolation, immediate measures to early confirmation of Covid-19 which was given the name of Hashmi-Asif Covid-19 formula for calculating early symptoms of Covid-19.

**Table.1.** Summary of Early common signs and symptoms appeared early onset of disease. All results are categorized and shown in percentages.

| Reference | Physical Symptoms |  | Mucus membrane |  | Blood Cells |  |  | Liver Enzymes |  |
| --- | --- | --- | --- | --- | --- | --- | --- | --- | --- |
|  | Fever | Cough | Dry | Hyperemic | Leucopenia | Lymphopenia | Thrombopenia | AST | ALT |
| W.Guanet al. | 43.8 | 67.8 | 33.7 | ~ | 33.7 | 87.2 | 36.2 | 22.2 | 21.3 |
| Lu X et al | 58.5 | 48.5 |  | 46.2 | 26.3 | 3.5 | ~ | 14.6 | 12.3 |
| Michelle et al | + | + | + | ~ | + | + | + | + | + |
| Haiyan et al | 36 | 19 | ~ | ~ | 19 | 36 | ~ | 8.3 | 5.5 |
| Haung C et al | 98 | 76 | ~ | ~ | 25 | 76 | 95 | 41.3 | + |
| Liu W et al | 100 | 100 | ~ | 83.7 | 66.7 | 100 | ~ | 66.7 | 16.6 |
| Y Song et al. | 1 | - | - | - | - | - | - | - | - |
| Wang et al, | 98.6 | 59.4 | ~ | ~ | + | 70.3 | + | + | + |
| Jin X, et al | 84.34 | 68.91 | 12.1<br>1 | ~ | + | + | + | - | - |
| Li Q et al. | + | - | - | - | + | + | - | - | - |
| + Present but not calculated<br>~ Not reported<br>- Normal values |  |  |  |  |  |  |  |  |  |

### Statistical Analysis

We investigated the relationship of frequent appearance of early sign and symptoms with diagnosed Covid-19 cases by Pearson and Spearman correlation coefficient (rho) two tail.^17^ Compiled data was analyzed statistically by using IBM SPSS Version.20.

## Results

W. Guan et al, described symptoms of more severe cases of covid-19 having fever in 975/1099 patients who were hospitalized with average temperature of 38.3 °C, having Leukopenia in 33.7% patients hospitalized for Covid-19, lymphocytopenia 87.2%, thrombocytopenia 36.2%, elevated Alanine aminotransferase 21.3% and elevated Aspartate aminotransferase were 22.2% and physician diagnosed pneumonia 91.1%. Cough 67.8%, fatigue 38.1% and sputum production in 33.7% in hospitalized patients.^6^ Result are compiled in Table.1. Case report of Covid-19 patient in USA described by Michelle L Holshue et al, provides the 15 days of hospitalization detailed course of disease of Covid-19 on daily observation of signs and symptoms. 35 year male presented with dry cough and fever experiencing from 3 days. Laboratory investigations showed Leukopenia and thrombocytopenia along with slight elevated liver enzymes AST and ALT on day 6^th^ and 7^th^ of illness respectively. Physical examination revealed dry mucus member while no symptoms or rhinorrhea and pneumonia were appeared before 9^th^ day of illness. Nausea and vomiting appeared on 4^th^ day of illness.^7^ Lu X et al. described detailed analysis of signs and symptoms of covid-19 in children. Lu showed leukopenia was 26.3%, in children hospitalized for Covid-19. Lymphocytopenia was 3.5%, increased ALT in 12.3%, and elevated AST 14.6%. Symptoms described as asymptomatic patients were 27/171, symptoms of upper respiratory tract infection were in 33/171 and symptoms of pneumonia were present in 111/171 of hospitalized children.^8^Now various publications explained concrete aspects of Covid-19 course of development and presentation. Longer incubation period (12.5 days) of SARS-Cov-2 embedded with some hidden advantages to halt from spreading epidemic and this requires wise judgments over understanding pathogenesis.^1^ Late appearance of primary symptoms of Covid-19 extends an opportunity^2^ to cessation of spreading and precluding from getting worsened and complicated by early detection and intervention. Course of Covid-19 followed concomitance to early symptoms appearance of mild temperature, cough, fatigue, nausea, vomiting, dry mucus membrane or hyperemic, dyspnea, consolidated pneumonia like lungs, accompanied with decline in blood oxygen saturation, leukopenia, lymphocytopenia, thrombocytopenia, elevation in Aspartate aminotransferase (AST), Alanine aminotransferase (ALT).^3-5^ Pediatric patients are difficult to prompt diagnosis and can remains asymptomatic up to 10 days.^9^ Haiyan Qu et al, prescribed epidemiological features of 36 children diagnosed with Covid-19 in Zheijiang, China. Haiyan observed clinical features of temperature in 36% children, cough in 19%, leucopenia in 19%, lymphopenia in 36% children diagnosed with Covid-19 on time of admission to hospital. AST was elevated in 8.3% children and ALT were elevated in 5.5% children on early stage of Covdi-19.^9^ Huang C et al, reported clinical features of hospitalized and laboratory confirmed Covid-19 cases in Wuhan, China. Huang C observed fever in 98%, Cough 76% in Covid-19 cases while blood investigations showed leucopenia 25%, lymphopenia 76%, thrombocytopenia 95% and elevated AST in 37%.^10^ Liu W et al, reported detection of 06 children with Covid-19 published in New England Journal of Medicine recorded fever (6/6) and cough (6/6) in all children diagnosed with Covid-19 under his study. Pharyngeal congestion was 83.7% (5/6), leucopenia (4/6) 66.7%, lymphopenia recorded in all children (6/6) 100%, platelets values were at lower limit <20×10^4^ in (3/5) half children. Elevated AST was (4/6) 66.3% and elevated ALT was 16.7 (1/6).^11^ Wang et al, in another publication appeared in JAMA described clinical symptoms of fever 98.6%, dry cough 59.4%, lymphopenia 70.3% in laboratory confirmed Covid-19, also described leucopenia, thrombocytopenia, elevated Alanine aminotransferase and aspartate aminotransferase in Covid-19 confirmed cases contrary Young et al, did not showed any laboratory findings in one study.^12-13^ Jin X et al, prescribed clinical features analyzed of 651 Covid-19 confirmed cases and find fever and cough with dry mucus membrane 84.34%, 68.91,% and 12.11% respectively. Leucopenia, lymphopenia and thrombopenia were also present but liver enzymes were in normal values.^14^ Detail features are prescribed in **Table.1**.

### Hashmi-Asif Covid-19 Chart

Sign and symptoms of Covid-19 can be classified into early symptoms and late symptoms. Early symptoms can be a point of consideration for getting early detection. Covid-19 diagnosis could be missed on early stage because of early symptoms being mild in nature but ambient to cater discreet evaluation for Covid-19 by calculating score on Hashmi-Asif Covid-19 chart as elaborated in Chart.1. Formula contains maximum of 28(10+18) scores out of which cumulative scoring ≥16/28 should be considered at high risk to be diagnosed with Covid-19, isolated immediately and should be evaluated by standard diagnostic procedure RT-PCR for SARS-Cov-2. The formula provides an easy approach to screen the suspects and carriers of Covid-19 earlier than previously being diagnosed. Blood oxygen saturation does not change much at early stages and the reason not included in calculation formula. Oxygen saturation decreases on advancing Covdi-19 and early time lapsed on taking such early measures. Decreased O_2_ gas in blood is critical situation to take urgent interventions. Hashmi-Asif covid-19 formula expedites health care providers in the developing countries lacking appropriate health facilities to access diagnosis of Covid-19. Hashmi-Asif Covid-19 formula based on the most common early presentations of the Covid-19. Separate calculation of scoring for sign and symptoms and blood biomarkers makes sensitivity for evaluating Covid-19. By using formula Covid-19 can be diagnosed approximately 3 days earlier than usual. It will provide an ample time to opt interventions for Covid-19 and to prevent intricate or reduced the mortality rate by early interventions.

### Statistical Results

Ten studies containing detailed information of 2282 (out of 2707) Covid-19 laboratory confirmed were analyzed for their common early sign and symptoms. And statistically pearsons correlation coefficient with rho measures were calculated by two tailed values for entirely each symptom. Each common symptomatic correlation with Covid-19 were statistically significant (sig.<0.01) except one symptoms of hyperemic mucus membrane. Fever 64.11% was significant Cough and dry mucus membrane values equally significant 0.000. leucopenia(19.06), lymphopenia (52.93) showed significance of 0.006 and 0.005 respectively. Thrombopenia (19.1) showed strong correlation (sig.0.013) with Covid-19 at sig. value (0.05). Elevated liver enzymes AST were presented at low values (12.79 & 11.34 respectively) showed strong correlation and statistically significant. All sign and symptoms cumulatively showed 29.5% sensitivity correlation among whole evaluated Covid-19. Data shows if all the confirmed cases were analyzed before confirmation with the early sign and symptoms 29.5% cases could be detected at and earlier than usual course of disease to be considered at very high risk to develop Covid-19. Statistical data is shown in **Table.2**.

**Table.2.** Symptomatic Correlation with early detection of high risk for development of Covid-19

| Symptoms | Total (2707)<br>2283 | Sig.(0.01) |
| --- | --- | --- |
| Fever | 1463(64.11%) | <0.001 |
| Cough | 1484(65%) | <0.000 |
| Dry Mucus Membrane | 449(19.67%) | <0.000 |
| Hyperemic Mucus<br>membrane | 85(3.72%) | <0.412 |
| Leucopenia | 435(19.06%) | <0.006 |
| Lymphopenia | 1208(52.93%) | <0.005 |
| Thrombopenia | 436(19.1%) | <0.013* |
| Elevated AST | 292(12.79%) | <0.007 |
| Elevated ALT | 259(11.34%) | <0.006 |
| *Sig. p value <0.05 |  |  |

## Discussion

SARS-Cov-2 is highly contagious, could spread rigorously like spread over the China mainland within 40 days infecting 72,314 cases during Covid-19 China epidemic. Zijian Fen et al, primarily described the epidemiological aspects of Covid-19, and asserted that Covid-19 has mild course of disease and mortality rate is 2.3%. According to Zijian Fen et al, many infected of mild and some severe cases survived the Covid-19 infection. Its symptoms are variable from mild to severe demanding assisted ventilation. 1.2% asymptomatic patients were confirmed by laboratory investigation. Many were suspected from their sign & symptoms and quarantined.^15^ Epidemic of Covid-19 in China attributed by spreading virus concomitant due to asymptomatic and late appearance of symptoms due to long incubation period.^3, 16^ Longer incubation period hides certain opportunity to get prepare and early prompt action against Covid-19. Asymptomatic cases may diagnose onset of disease and earlier symptoms appearing during course of disease also holds credible opportunity to halt earlier than usual diagnosis and intervention. Early detection could only be possible b assessing sign and symptoms like evaluated from various studies. Evaluation of collected data provides important clinical features which could provide comprehensive and reliable information for suspecting cases to develop Covid-19. Among various studied sign and symptoms only those were considered in this evaluation those were highly correlated with development of Covid-19. Fever of mild to moderate grade was present 64.11% and cough has highest correlation with development of Covid-19 appeared in the study. Cough was present in 65% confirmed cases of Covide-19. However, asymptomatic cases developed without early signs are hard to be detected before progressed Covid-19. Coughing enormously present during course of disease and could of dry or productive in nature, may accompanied with secondary to former infection or underlying pathology. Dry cough or dry mucus membrane holds significant correlation^7^ with development of Covid-19. Leukopenia and thrombopenia is characteristic feature of Covid-19 infection and can be assessed before onset of symptoms. Elevated AST and ALT also hold substantial relation with Covid-19.

### Significance of Hashmi-Asif Covid-19 Formula

Early detection for Covid-19 in symptomatic cases showed its sensitivity and could isolate Covid-19 case at an early stage. Calculation chart is given name of Hashmi-Asif Chart for early detection of Covid-19, given in Chart.1. Formula contains maximum of 28(10+18) scores out of which cumulative scoring ≥16/28 should be considered at high risk to be diagnosed with Covid-19, isolated immediately and should be evaluated by standard diagnostic procedure RT-PCR for SARS-Cov-2. The formula provides an easy approach to screen the suspects and carriers of Covid-19 earlier than previously being diagnosed. Hashmi-Asif covid-19 formula expedites health care providers in the developing countries lacking appropriate health facilities to access diagnosis of Covid-19. Hashmi-Asif Covid-19 formula based on the most common early presentations of the Covid-19. Separate calculation of scoring for sign and symptoms and blood biomarkers makes sensitivity for evaluating Covid-19. By using formula Covid-19 can be diagnosed approximately 3 days earlier than usual. It will provide an ample time to opt interventions for Covid-19 and to prevent intricate or reduced the mortality rate by early interventions.

## Conclusion

Studies showed a strong correlation of early sign and symptoms leading to development of Covid-19. Sensitivity and specificity of these symptoms holds potential 0f 29.5% chances to detect earlier progression to develop Covid-19. By utilizing these symptoms into a calculated chart is an easy way to get suspects developing Covid-19 and taking early confirmation from nasopharyngeal and oro-pharyngeal swab detection by RT-PCR. Scoring chart expedite early detection and intervention consequence to reduced morbidity time and prevent from progression of disease to severe condition. Symptomatic appearance among Covid-19 infected cases Hashmi-Asif Covid-19 chart hold sensitivity of 95% to early detection which will surely reduce spreading spectrum of Covid-19 to prevent epidemic out break or slow downed its spread. Early detection chart will make an easy approach to manage Covid-19 cases.

## Data Availability

Available data on early sign and symptoms of Covid-19 on World Health Organization (WHO) webpages, New England Journal of Medicine (NEJM), Nature Journal, Journal of American Medical Association (JAMA) network, British Medical Journal (BJM), Lancet

---

**Chart.1:**
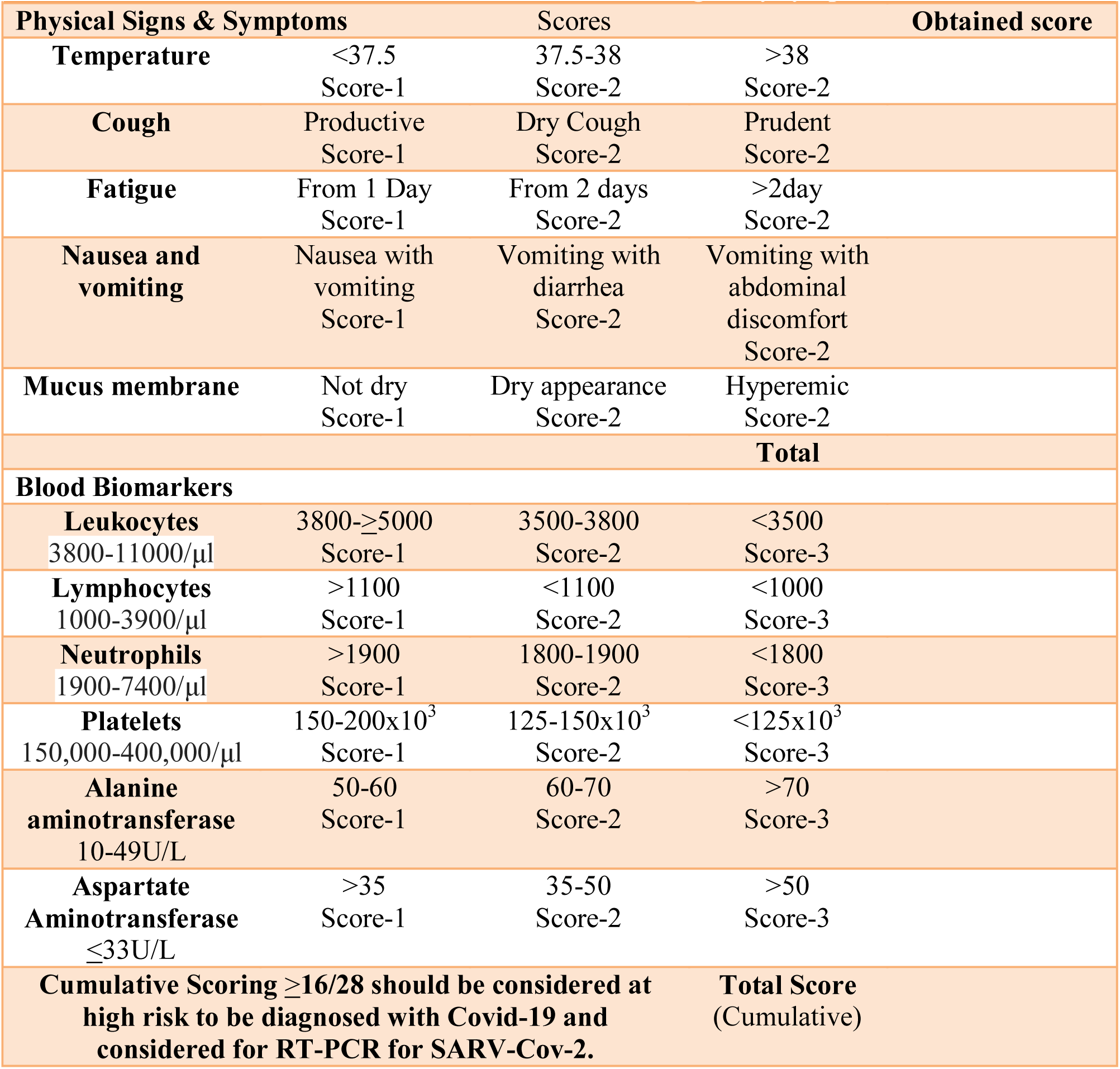
Hashmi-Asif Covid-19 Chart for calculating early symptoms of Covid-19

## Declaration

Authors declared no conflict of interest about this manuscript.

## References

1. Joseph T. Wu et al, Estimating clinical severity of Covid-19 from the transmission dynamics in Wuhan, China, Nature Medicine March 19, (2020) https://doi.org/10.1038/s41591-020-0822-7

2. Na Zhu et al, A Noval Corona Virus form Patient of Pneumonia in China, 2019. N Engl J Med 2020;382:727–33

3. David M. Morens et al. Escaping Pandora’s Box — Another Novel Coronavirus, N Engl J Med published on February 26,2020, at NEJM.org. DOI: 10.1056/NEJMp2002106

4. Li Q, Guan X, Wu P, et al. Early transmission dynamics in Wuhan, China, of novel coronavirus–infected pneumonia. N Engl J Med. DOI:10.1056/NEJMoa2001316

5. Anthony S. Fauci et al, Covid-19-Navigating the uncharted, editorial published on February 28, 2020, at NEJM.org DOI: 10.1056/NEJMe2002387

6. W. Guan et al. Clinical Characteristics of Coronavirus Disease 2019 in China published on February 28,2020, at NEJM.org. DOI: 10.1056/NEJMoa2002032

7. Michelle L. Holshue et al, First Case of 2019 Novel Coronavirus in the United States N Engl J Med 2020;382:929–36.

8. Lu X, Zhang L, Du H, et al. SARS-CoV-2 infection in children. N Engl J Med. DOI: 10.1056/NEJMc2005073.

9. Haiyan Qiu, Junhua Wu, Liang Hong, Yunling Luo, Qifa Song, Dong ChenClinical and epidemiological features of 36 children with coronavirus disease 2019 (COVID-19) in Zhejiang, China:an observational cohort study Lancet Infect Dis 2020 Published Online March 25, 2020https://doi.org/10.1016/S1473-3099(20)30198-5

10. Huang C, Wang Y, Li X, Ren L, Zhao J, Hu Y, Zhang L, Fan G, Xu J, Gu X, Cheng Z, Yu T, Xia J, Wei Y, Wu W, Xie X, Yin W, Li H, Liu M, Xiao Y, Gao H, Guo L, Xie J, Wang G, Jiang R, Gao Z, Jin Q, Wang J, Cao B. Clinical features of patients infected with 2019 novel coronavirus in Wuhan, China. Lancet. 2020 Feb 15;395(10223) 497–506

11. Liu W, Zhang Q, Chen J, et al. Detection of Covid-19 in children in early January 2020 in Wuhan, China. N Engl J Med. DOI: 10.1056/NEJMc2003717

12. Y Song, P Liu, X L Shi, Y L Chu, J Zhang, J Xia, X Z Gao, T Qu, M Y WangSARS-CoV-2 induced diarrhoea as onset symptom in patient with COVID-19 Gut Mar 2020, gutjnl-2020-320891; DOI: 10.1136/gutjnl-2020-320891

13. Wang D, Hu B, Hu C, et al. Clinical characteristics of 138 hospitalized patients with 2019 novel coronavirus-infected pneumonia in Wuhan, China. JAMA 2020.doi:10.1001/jama.2020.1585.

14. Jin X, et al. Epidemiological, clinical and virological characteristics of 74 cases of coronavirus-infected disease 2019 (COVID-19) with gastrointestinal symptoms Gut 2020;0:1–8. doi:10.1136/gutjnl-2020-320926

15. Zijian Fen et al, Novel Coronavirus Pneumonia Emergency Response Epidemiology Team. Vital surveillances: the epidemiological characteristics of an outbreak of 2019 novel coronavirus diseases (COVID-19)—China, 2020. China CDC Weekly. Accessed February 20, 2020. http://weekly.chinacdc.cn/en/article/id/e53946e2-c6c4-41e9-9a9b-fea8db1a8f51

16. D. Chang, M. Lin, L. Wei, L. Xie, G. Zhu, C.S. Dela Cruz, et al. Epidemiologic and clinical characteristics of novel coronavirus infections involving 13 patients outside Wuhan, China JAMA (2020 Feb 7), 10.1001/jama.2020.1623

17. Gregory A. Roth et al, Demographic and Epidemiologic Drivers of Global Cardiovascular Mortality N Engl J Med 2015;372:1333–41. DOI: 10.1056/NEJMoa1406656

